# The Impact of Artificial Intelligence on the Health Economy, Workforce Productivity, and Administrative Efficiency: A Systematic Review

**DOI:** 10.1101/2025.10.05.25337345

**Authors:** John Tayu Lee, Valerie Tzu Ning Liu, Shehzad Ali, Pau-Chung Chen, Chien-Chang Lee, Chang-Wei Huang, Vincent Cheng-Sheng Li, Hsiao-Hui Chen, Wei-Jou Duh, Tiara Marthias, Rifat Atun

## Abstract

**Background:** Healthcare systems globally are under increasing financial and operational strain due to aging populations, rising expenditures, and workforce shortages. Amid these challenges, artificial intelligence (AI) has emerged as a promising tool to enhance system-level performance, particularly in cost reduction, productivity gains, and administrative efficiency.

**Objective:** This review aims to synthesize existing evidence on the macro and system-level impact of AI implementation across three key domains: the health economy, workforce productivity, and administrative efficiency.

**Methods:** A systematic review methodology was employed, allowing for the integration of diverse data sources and study types. Literature was systematically identified through PubMed and Google, covering publications from 2020 to July 2025. Studies were included if they evaluated AI’s impact on national or regional health expenditure, labor restructuring, or process efficiency. Thematic synthesis was guided by a conceptual framework modeling AI as a system-wide catalyst.

**Results:** Twenty-four studies were included. AI implementation demonstrated potential cost savings of 5–10% in national health expenditures, driven by automation in hospital operations and administrative processes. AI-supported interventions reduced diagnostic time by up to 90% and treatment costs by over 30% in specific applications, such as cancer diagnosis and radiotherapy. Administrative tools, including AI-assisted documentation and claims processing, achieved efficiency gains of up to 40%. However, reliance on simulated models, short-term studies, and single-center data limits generalizability.

**Conclusions:** AI presents significant potential to enhance health system efficiency and reduce costs. Real-world implementation studies, standardized outcome metrics, and robust governance frameworks are essential to validate these gains and ensure equitable, sustainable adoption.

## INTRODUCTION

Health systems around the world are facing growing pressure. Spending on healthcare is rising faster than many economies can keep up with due to rapid aging population^1^. The percentage of GDP spent on health has shown a significant increase over the past decades. According to projections, healthcare spending in the United States is expected to grow faster that the GDP, reaching 19.7% of GDP by 2032, up from 17.3% in 2022^2^. This trend reflects a broader global pattern where health spending as a share of GDP has been increasing, driven by factors such as economic development, demographic changes, and policy shifts. For instance, global health spending was 8.6% of the global economy in 2016 and is projected to reach 9.4% by 2050 ^3^.

The escalating cost occurs while many health systems struggling with insufficient healthcare funding from both the public and private sector. For example, the UK’s National Health Service (NHS) faces a projects £ 4.8 bill shortfall in its 2024/2025 revenue budget, raising concerns about potential service cuts without additional funding ^4^. In the United States, proposed federal cuts of $ 880 to Medicaid over the next decade threaten to reduce access to essential services cater for vulnerable populations ^5^.

Additionally, many health system has shortages in the healthcare workforce, making it challenging to meet the growing demand^6^. This issue is compounded by the maldistribution of healthcare professional across geographical area, leading to inequalities in service provision in underserved and remote areas. Adding to this burden is the increasing amount of administrative work that clinicians and managers face growing administrative burden, diverting valuable time from direct patient care. According to a time and motion study, physicians in ambulatory practice spend 49.2% of their time on electronic health record (EHR) and desk work and 27.0% on direct clinical face time with patients during office hours^7^. Another study in the Journal of General Internal Medicine found that physicians spend 20.7% of their time on EHR input alone and 7.7% on administrative activities, with 66.5% of their time on direct patient care (including multitasking with EHR) ^8^.

Against this backdrop, Artificial Intelligence (AI) and Machine Learning (ML) have captured attention as tools that could potentially reshape how health systems function. AI is not new—it began with early attempts to simulate human thinking using symbolic systems and rule-based expert programs in the 1980s. These tools could be useful in narrow areas but were limited by their inability to handle complexity and real-world uncertainty. What has changed in recent years is the rise of new forms of AI, particularly deep learning and large-scale foundation models. These systems learn from enormous amounts of data and can now perform tasks that once required human intelligence—such as reading radiology scans, understanding natural language, writing computer code, or even engaging in meaningful conversation. Tools like GPT and Gemini represent a leap forward not only in accuracy but also in general intelligence. In healthcare, they are already being tested for clinical decision support, predictive risk modelling, and automating administrative tasks. At present, AI tools have been increasingly used and accepted in clinical practice, particularly in imaging and diagnosis. LLMs have also been used for documentation, administrative, or coding work in clinical settings ^9,10^.

But alongside this rapid progress, concerns remain. These systems rely heavily on the data used to train them—data that may be incomplete, biased, or not representative. AI decisions can also be difficult to interpret or explain, which raises issues of trust, accountability, and ethics ^11^. While much attention has focused on AI’s technical capabilities, its broader impact on health systems as a whole requires deeper investigation. From a policy prospective, it is essential to examine the macro-level impact of AI on health economy. This study reviews the current evidence on how AI is influencing three key areas: (1) overall health spending and cost control, (2) administrative efficiency, and (3) workforce productivity.

## RESULTS

### Summary of Included Studies

This review includes 24 studies examining the macro-level impact of artificial intelligence (AI) across three key domains: health economy, workforce productivity, and administrative efficiency. Of these, 9 studies were conducted in the United States, and the remainder spanned global or multi-country settings, including Taiwan, Japan, Bengal articles^12-19^.

Medical imaging emerged as the most common application area, with nine studies focusing on AI’s role in interpreting radiologic, endoscopic, or pathological images ^15,16,20,21^. These studies investigated tasks such as detection^15,16,22^, diagnosis^16,22-24^, reduction of interpretation time ^15,16^, and workflow optimization ^15,16,23^.

Nine studies^12,14,17-20,22,24,25^explicitly addressed the health economic impact of AI, including cost savings^12-14,19,20,24^, budget impact^19,20,24^, and reduction in treatment costs^14^. Administrative efficiency was the focus of three studies ^13,26^, each addressing the potential of AI to automate and streamline non-clinical tasks such as clinical documentation ^13,18^, and claims processing ^13,19^.

**Table 1.** Summary characteristics of included studies.

| Study | Type | Country | Data | Intervention of Interest |
| --- | --- | --- | --- | --- |
| **Abramoff et al<sup>21</sup>** | Workforce productivity | Bengal | A clinic with a very large demand , Deep Eye Care Foundation (DECF) in Bangladesh | LumineticsCore (IDx-DR)<br>Diabetic eye exams. |
| **Natesan et al<sup>12</sup>** | Health economy | The USA | Duke Cancer Center<br>Duke Health System<br>cost accounting system, Medicare, and Medicaid | ML(XGBoost)-identified high-risk courses (≥10% risk of acute care during RT). |
| **Lavoie et al<sup>13</sup>** | Administrative Efficiency | The USA | Analysis of existing US healthcare spending data and administrative costs. | Application of AI tools to automate and streamline non-clinical administrative processes. |
| **Sahni et al**<sup>19</sup> | Health economy | The USA | Expert inputs, McKinsey internal experiments, and published research. National Health Expenditures data (2019) | (ML) applications in healthcare ,(NLP) is used to extract text from clinician notes for documentation or coding. |
| **Kacew et al**<sup>14</sup> | Health economy | The USA | Secondary research (peer-reviewed literature and Medicare data) | AI for Cancer Diagnosis Cost. |
| **Yacoub et al**<sup>15</sup> | Workforce productivity | The USA | Medical University of South Carolina | AI-Rad Companion. |
| **Hunter et al**<sup>16</sup> | Workforce productivity | The USA | Picture archiving and communication system (PACS) single-view CXRs of adult inpatient. | The diagnostic accuracy and impact on radiologist report turnaround times of an FDA-approved AI tool for pneumothorax (PTx) detection on inpatient chest X-rays (CXR) in our institution's radiology practice at a large academic medical center |
| **Worrall et al**<sup>22</sup> | Health economy | The USA | Medicare. | AI supports pharmacist-led medication adherence programs. |
| **McKinsey Company et al 2023**<sup>17</sup> | Health economy | Global | Extensive global data and expert insights | Generative AI-Product R&D software engineering, Customer documentation generation, Generate content for commercial representatives, Contract generation. |
| **McKinsey Company et al 2020**<sup>18</sup> | Health economy<br>Workforce productivity | Global | Extensive global data and expert insights | This definition is deliberately broad; it includes the application of rules-based systems |
|  | Administrative Efficiency |  |  | as well as cutting-edge methodologies that include classic machine learning, representation learning and deep learning. Examples of AI applications would include NLP, image analysis and predictive analytics based on machine learning. |
| **Mori et al**<sup>20</sup> | Health economy | Japan | Large-scale single-center clinical trial | EndoBRAIN |
| **Dembrower et al**<sup>27</sup> | Workforce productivity | Sweden | Karolinska University Hospital | Ai based triaging of breast cancer screening mammograms. |
| **Rozario et al**<sup>26</sup> | Administrative Efficiency | Canada | Oakville Trafalgar Memorial Hospital | Python programming language combined with the open source OR-Tools software suite from Google AI. |
| **Willmen et al**<sup>24</sup> | Health economy | Germany | Hannover Medical School. | A Diagnostic Decision Support System (DDSS) is an artificial intelligence–based system that supports physicians in diagnosing rare diseases. |
| **Lauritzen et al**<sup>28</sup> | Workforce productivity | Danish | A single European institution. | Transpara employs deep convolutional neural network (DCNN) technology to detect suspicious breast cancer lesions on full-field digital mammography. |
| **Tong et al**<sup>29</sup> | Workforce productivity | China | First Affiliated Hospital of Sun Yat-sen University and the Third Affiliated. | A deep learning AI model assists in improving the diagnostic accuracy of |
|  |  |  | Hospital of Sun Yat-sen University. | thyroid nodules by enhancing the performance of ultrasonographic diagnosis. |
| **Brix et al<sup>25</sup>** | Health economy | Finland | Hospital's radiology information system (RIS). | Deep Resolve Boost enhances MRI scanner throughput and significantly shortens MRI scan time while maintaining or improving image quality. |
| **Chen et al<sup>30</sup>** | Workforce productivity | Taiwan | Chi Mei Medical Center. | A+ Nurse is a ChatGPT-based large language model (LLM) tool that enhances the efficiency and accuracy of nursing documentation. |
| **Ahsen et al<sup>31</sup>** | Workforce productivity | Korea | A tertiary hospital in South Korea. | A commercially available DLD. The algorithm was designed to identify suspicious areas for 9 abnormal radiologic findings. |
| **Almagharbeh et al<sup>32</sup>** | Workforce productivity | Jordan | Two governmental hospitals and three private hospitals in Amman, Jordan. | AI-based decision support systems (DSS) on the operational processes of nurses in critical care units (CCU). |
| **Evans et al<sup>33</sup>** | Administrative Efficiency | Australia | One allied health organization. | Lyrebird Health is an AI medical scribe. |
| **Hadi et al<sup>34</sup>** | Workforce productivity | Saudi Arabia | The radiology department of a tertiary referral hospital in Makkah, Saudi Arabia. | ChatGPT-4o. |
| **McKinney et al<sup>35</sup>** | Workforce productivity | the UK and the USA | UK National Health Service Breast Screening Programme and Northwestern | AI system that is capable of surpassing human experts in breast cancer prediction. |
|  |  |  | Memorial Hospital<br>(Chicago). |  |
| Kyono et al<sup>36</sup> | Workforce<br>productivity | the UK | Six UK National<br>Health Service Breast<br>Screening Program<br>centers. | The Autonomous<br>Radiologist Assistant<br>(AURA), integrating ML,<br>CNN, DNN, and a<br>hybridized approach, is<br>applied to breast<br>cancer screening. |

#### Impact of Health Economy or Systems-Level Spending

The study, conducted by a team of researchers from Harvard, offers one of the most comprehensive assessments of the potential financial impact of AI in healthcare sector^19^. It estimates that over the next five years, AI could generate annual net savings of $200 billion to $360 billion in the U.S. healthcare sector without compromising quality or access, based on 2019 dollars. This represents a 5% to 10% reduction in total U.S. healthcare spending. The estimate is based on data from the National Health Expenditure, specifically total hospital revenues and total hospital costs^19^.

Complementing these quantitative projections, the EIT Health & McKinsey report highlights AI’s systemic role in sustaining healthcare delivery under financial and workforce pressures, particularly in Europe, by enhancing productivity and enabling more proactive models of care^18^. At a broader economic frontier, McKinsey’s assessment of generative AI estimates an annual global value creation of USD 2.6–4.4 trillion, with USD 60–110 billion accruing specifically to the pharmaceutical and medical-product industries, pointing to further downstream effects on healthcare affordability and innovation^17^.

#### Costs Reduction or Saving for Treatment

Few studies identifying cost savings associated with the use of AI. s. One example is provided by Mori et al., who analyzed AI-aided colonoscopy for polyp diagnosis. They compared traditional methods with an AI-aided approach for detecting diminutive colorectal polyps. Their findings indicated reductions in average colonoscopy costs of approximately 18.9% in Japan, 6.9% in England, 7.6% in Norway, and 10.9% in the United States. Thus, the use of AI results in substantial cost savings for colonoscopy^20^. A study on metastatic colorectal cancer demonstrates that deploying high-sensitivity AI for MMR/MSI status determination, followed by confirmatory testing for negative cases, resulted in projected savings of $400 million— approximately 12.9% of total diagnostic and treatment costs—compared to next-generation sequencing alone ^14^. Another cost analysis focusing on machine learning-guided radiotherapy interventions showed a statistically significant reduction in acute care visits and a halving of treatment costs per course ($1,494 vs. $3,110), suggesting strong cost-efficiency when targeting high-risk cases using AI triage ^12,14^.

Additionally, a retrospective study showed substantial cost reductions through improved medication adherence and patient outcomes using AI. Findings indicated that after the introduction of a pharmacist-led, AI-supported program, medication adherence improved across all disease measures. Importantly, adherent patients demonstrated notable reductions in overall healthcare expenditures compared to nonadherent patients, with cost savings of 31% for hypertension, 25% for hyperlipidemia, and 32% for diabetes. However, external validity may be limited by claim data constraints, potentially impacting the reflection of actual clinical outcomes due to missing or inaccurate data^22^.

#### Efficiency Gains in Medical Imaging

In radiology, the implementation of an AI tool for pneumothorax detection was shown to significantly improve report turnaround time, reflecting measurable gains in workflow efficiency16. Similarly, a prospective randomized study found that integrating an automated AI platform into chest CT interpretation reduced reading time by 22.1%, demonstrating AI’s potential to enhance radiologist productivity15. Similarly, in breast cancer screening, an international evaluation published in *Nature* demonstrated that an AI system outperformed radiologists in both the US and UK, reducing false positives by up to 5.7% and false negatives by 9.4%, while cutting second-reader workload by 88%^37^.

These findings align with *Transforming Healthcare with AI*, which identifies diagnostic speed and accuracy as key areas where AI is already delivering impact. The report emphasizes that imaging-heavy specialties such as radiology, pathology, and ophthalmology are being actively reshaped by AI, describing imaging applications as “low-hanging fruit” due to their readiness to provide immediate, tangible benefits in clinical settings^18^.

#### Efficiency Gains in Medical Documentation

One study identified clinical documentation as a significant component of administrative costs and discussed how tools such as NLP can extract relevant data from clinical notes. This can help ease documentation demands from payers and accelerate claims approvals, emphasizing the link between documentation processes and administrative efficiency^13^.

Similarly, Analysis by McKinsey group notes that AI can take on administrative tasks, including the use of NLP to process clinical notes—highlighting its role in automating clinical documentation and improving workflow efficiency in this area^18^.

Recent empirical work further illustrates these benefits in practice. Rozario and colleagues demonstrated that applying a machine-learning optimization model to operating room scheduling significantly reduced overtime by 21%, translating to an estimated cost saving of USD 469,000 over three years^26^. In allied health private practice, Evans et al. found that using an AI scribe decreased time spent on clinical notes and letters, reduced after-hours documentation, and improved both productivity and therapeutic alliance between clinicians and patients^33^. Together, these findings highlight AI’s dual potential: streamlining administrative processes while simultaneously enhancing the efficiency and quality of clinical workflows.

#### Efficiency Gains in Claim Management

In terms of administrative efficiency, AI tools have shown some of the clearest and most immediate benefits^13,18,19^. Studies suggest that automating claims processing, prior authorization, quality assurance, and credentialing could eliminate as much as $168 billion in annual U.S. administrative spending^13^. AI applications in this area include centralized digital claims clearinghouses, NLP-based documentation extraction, and fully electronic prior authorization systems^13^. For instance, centralizing claims processing was estimated to save $300 million annually, while electronic authorization pipelines could yield an additional $417 million in savings. The McKinsey and NBER analyses both highlight administrative automation as a primary driver of system-level savings, estimating that 40–50% of AI-derived cost reductions for hospitals and physician groups stem from back-office and billing functions.

Potential in Claims Management and Member Services: AI could lead to significant savings by addressing repetitive administrative processes such as processing prior authorisations or adjudicating claims. AI-enabled identification models can streamline operations and inform efforts to reduce fraud, waste, and abuse^19^.

Both McKinsey’s *Reports* (2020, 2023) highlight AI’s transformative potential but differ in scope: the former focuses on AI’s role in improving healthcare delivery and reducing system waste, while the latter explores cross-industry automation of knowledge work. Both reports also address labor implications—*The Economic Potential of Generative AI* (2020) points to potential displacement of high-skill tasks, whereas *Transforming Healthcare with AI* emphasizes relieving administrative burdens in healthcare and the importance of digital upskilling. While *Generative AI* focuses on theoretical value creation, *Transforming Healthcare with AI* ties economic gains to specific healthcare efficiencies and underscores the need for governance, funding, and workforce readiness to realize AI’s full potential^18,38^.

In summary, Figure 2 illustrates a mechanism-based framework that was consistently described across several studies included in this review. Beginning with inputs in the form of AI technologies—including applications in natural language processing, image analysis, and machine learning—the framework highlights the key mechanisms through which these technologies enact change. These include process automation, decision support, and resource optimization. As these mechanisms are embedded into healthcare operations, they contribute to measurable improvements in productivity, efficiency, and service quality. These system-level outcomes, in turn, drive economic impact through cost savings, improved health outcomes, and better use of resources. This pathway emphasizes that the economic benefits of AI are not simply a function of the technology itself, but rather emerge from its integration into and influence on health system processes.

**Figure 1.**
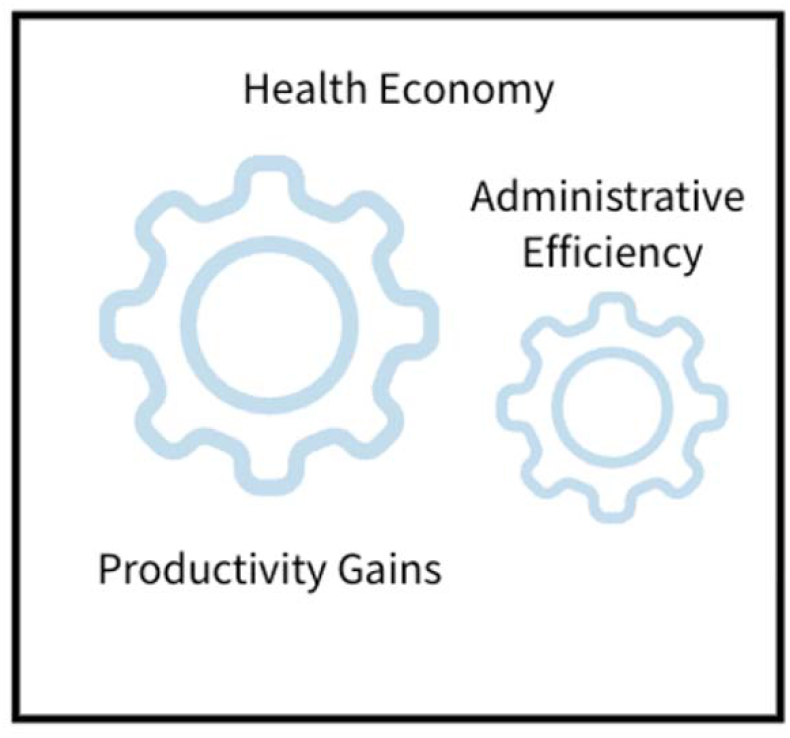
Conceptual framework of the impact of Al on health economy.

**Figure 2.**
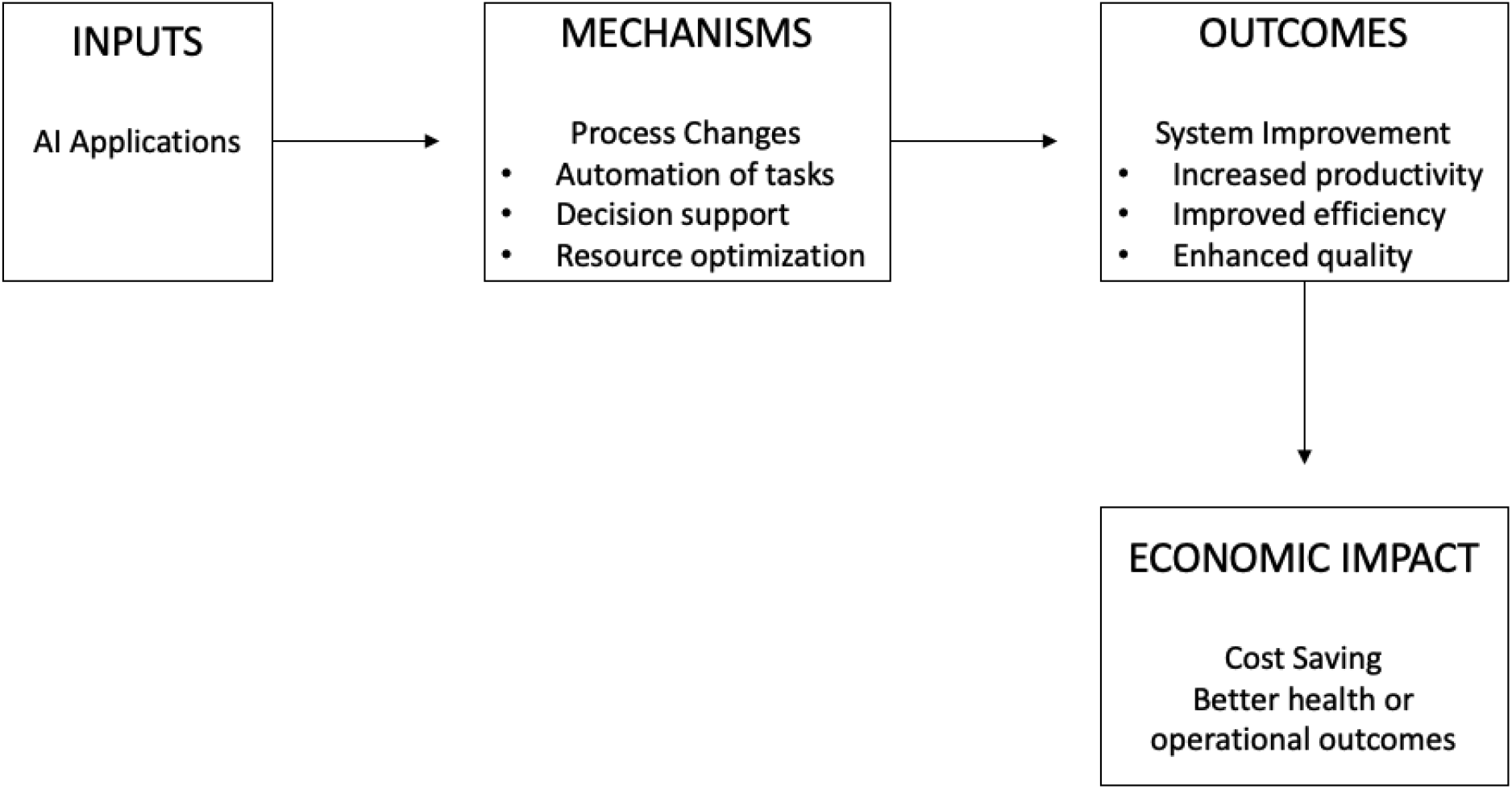
Mechanism of AI application impact health economy

**Table 2.** Methodological Approaches for Examining Health Economy Outcomes.

| Study | Measurement | Statistical Method | Findings | Limitations |
| --- | --- | --- | --- | --- |
| **Natesan et al<sup>12</sup>** | The primary outcome was difference in mean total medical costs during and 15 days after RT. | A cost-minimization analysis was conducted to compare the standard treatment strategy with the intervention approach. randomized controlled study. | Significant reduction in total healthcare costs: After adjusting for patient and disease factors, the average total healthcare cost per treatment course was significantly lower in the intervention group at \$1,494, compared to \$3,110 in the standard care group (P = 0.03). | Nonfinancial benefits, such as improvements in patient health, were not considered. A 10% risk threshold for intervention was selected based on available personnel resources. |
| **Kacew et al<sup>14</sup>** | Total costs of testing and first-line therapy across different diagnostic strategies for determining MMR/MSI status in metastatic colorectal cancer. | Financial and clinical modelling to compare costs of different diagnostic algorithms (NGS alone, PCR/IHC panel alone, AI-based strategies with and without confirmatory tests). | The testing strategy that resulted in the greatest project cost savings (including testing and first-line drug cost) compared to next-generation sequencing alone in newly-diagnosed metastatic colorectal cancer was using high-sensitivity artificial intelligence followed by confirmatory high-specificity polymerase chain reaction or immunohistochemistry panel for patients testing negative by artificial intelligence (\$400 million, 12.9%). | The recent study used theoretical models, and its results need to be validated using real-world data. • The accuracy of the estimates depends on the validity of the various sources used to establish the model input assumptions. |
| **Sahni et al<sup>19</sup>** | Potential net savings opportunity across | Estimate a gross savings percentage for each | AI adoption within the next five years could result in savings of 5 to 10 percent of health | The insights are not based on randomized control trials or quasi-experimental evidence. |
| | different healthcare stakeholders by using existing AI technologies | domain. Breaking down the industry into five stakeholder groups. | care spending, or \$200 billion to \$360 billion annually in 2019 dollars | Focus only on what is possible using existing technologies. |
| **Worrall et al<sup>22</sup>** | Medication Adherence Measures Disease Control Measures Overall health care expenditures (cost saving) | Retrospective, quasi-experimental analysis. | A reduction of \$3388 per member per year and \$282 per member per month in total cost of care. | Claims Data may be inaccurate, limited ability to reflect actual clinical outcomes, limited representativeness. Only an approximation of actual medication-taking behavior. |
| **McKinsey Company et al 2023<sup>17</sup>** | Economic Potential R&D Efficiency | To assess the potential value of generative AI, they updated a proprietary McKinsey database of potential AI use cases and drew on the experience of more than 100 experts in industries and their business functions. | Generative AI could have a significant impact on the pharmaceutical and medical-product industries—from \$60 billion to \$110 billion annually. Improve automation of preliminary screening. | The need for a human in the loop Explainability Privacy considerations |
| **Mori<sup>20</sup>** | cost per colonoscopy and gross annual reimbursement for colonoscopies | Average costs per colonoscopy based on the worst-case scenario. | In Japan the average colonoscopy cost under the diagnose-and-leave strategy (supported by AI prediction) was US\$119 lower, The expected gross annual reimbursement savings for colonoscopies would be US\$149,220,133. | Single-center study Potential overestimation of AI performance |
| **McKinsey Company et al 2020<sup>18</sup>** | A comprehensive understanding of AI's impact in healthcare, encompassing multiple dimensions including technology, economics, workforce, regulation, and user experience. | Qualitative and Quantitative Analysis. | AI has tremendous potential to transform healthcare by improving care outcomes, patient experience, and access to medical services, while also enhancing efficiency. | N/A |
| **Willmen et al<sup>24</sup>** | Costs | Descriptive statistics. | DDSS can achieve up to a 50% cost reduction by avoiding unnecessary duplicate tests and by enabling faster diagnosis. | The study employed a retrospective approach, and the results are exploratory in nature. Case scope limitation: The patient cases examined represent only a small subset of specific rare diseases; therefore, the findings cannot be generalized to all rare diseases. |
| **Brix et al<sup>25</sup>** | Annual total cost savings. | Comparative analysis of financial feasibility. | This investment is expected to save €399,000 annually in MRI service costs. | The revenue changes resulting from the increase in MRI examinations cannot be directly generalized to other regions. |

**Table 3.** Methodological Approaches for Examining Administrative Efficiency.

| Study | Measurement | Statistical Method | Findings | Limitations |
| --- | --- | --- | --- | --- |
| **Lavoie et al<sup>13</sup>** | Potential for cost savings by | Not clear. | Adoption of these tools can eliminate \$168 | Assessing administrative |
|  | leveraging AI to streamline billing, insurance, credentialing, and quality assurance processes. |  | billion in annual administrative costs. | costs is difficult given the limited transparency and lack of standardized reporting practices. |
| **Rozario et al<sup>26</sup>** | Overtime frequency, Undertime frequency, Overtime minutes and used Overtime cost. | Assessed retroactively. | Overtime reduced by 20.99%, Undertime reduced by 19.03% and On-time increased to 55%. | Requires programming skills to implement. |
| **Evans et al<sup>33</sup>** | Productivity and percentage of time AHPs reported using the AI scribe. | Inferential Statistics. | saving approximately 17-min per day on clinical notes and 9-min per day on letters. Productivity increased by an average of 5.8 %. | All allied health professional (AHP) participants were recruited from a single organization, which may limit the generalizability of the findings. Potential sampling bias. |

**Table 4.**
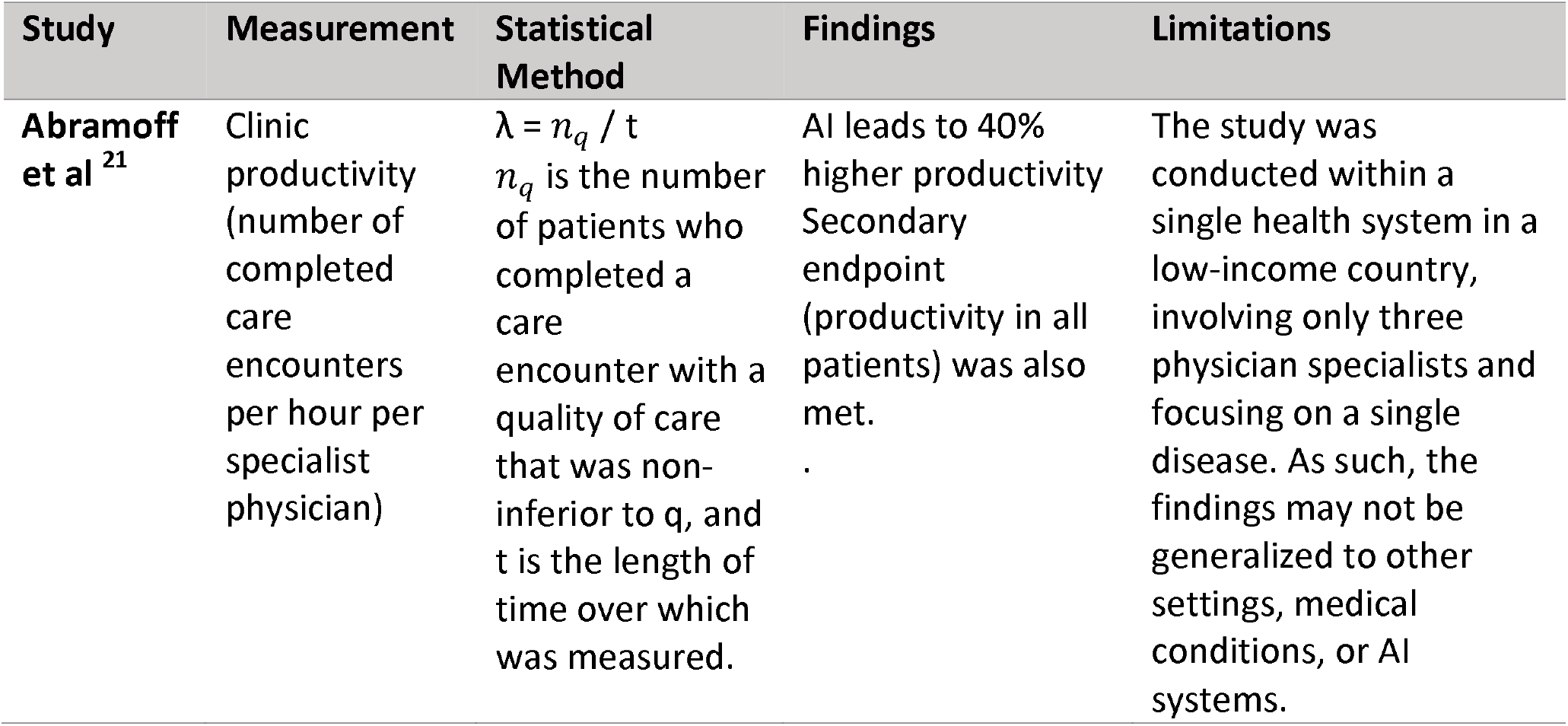
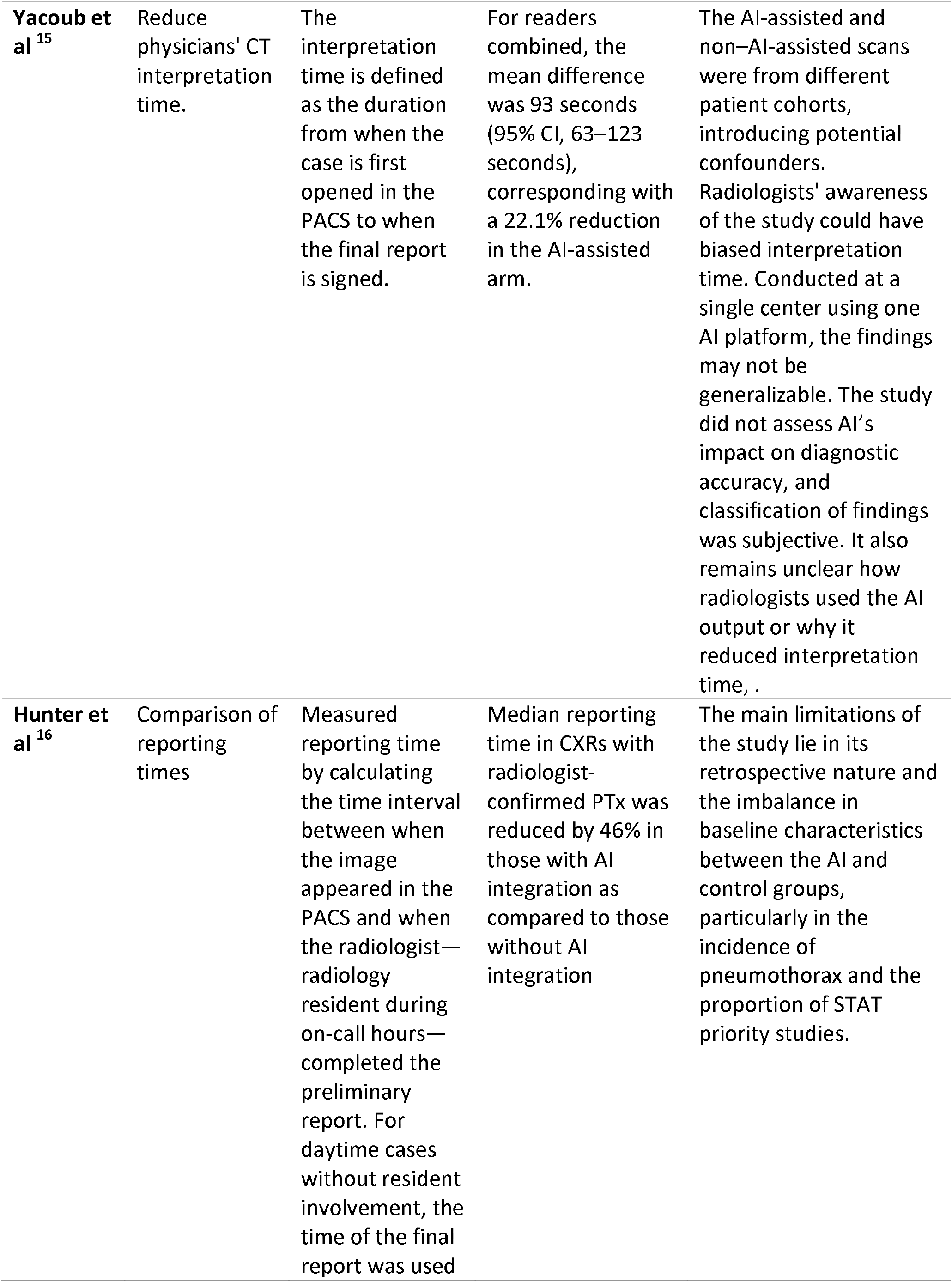
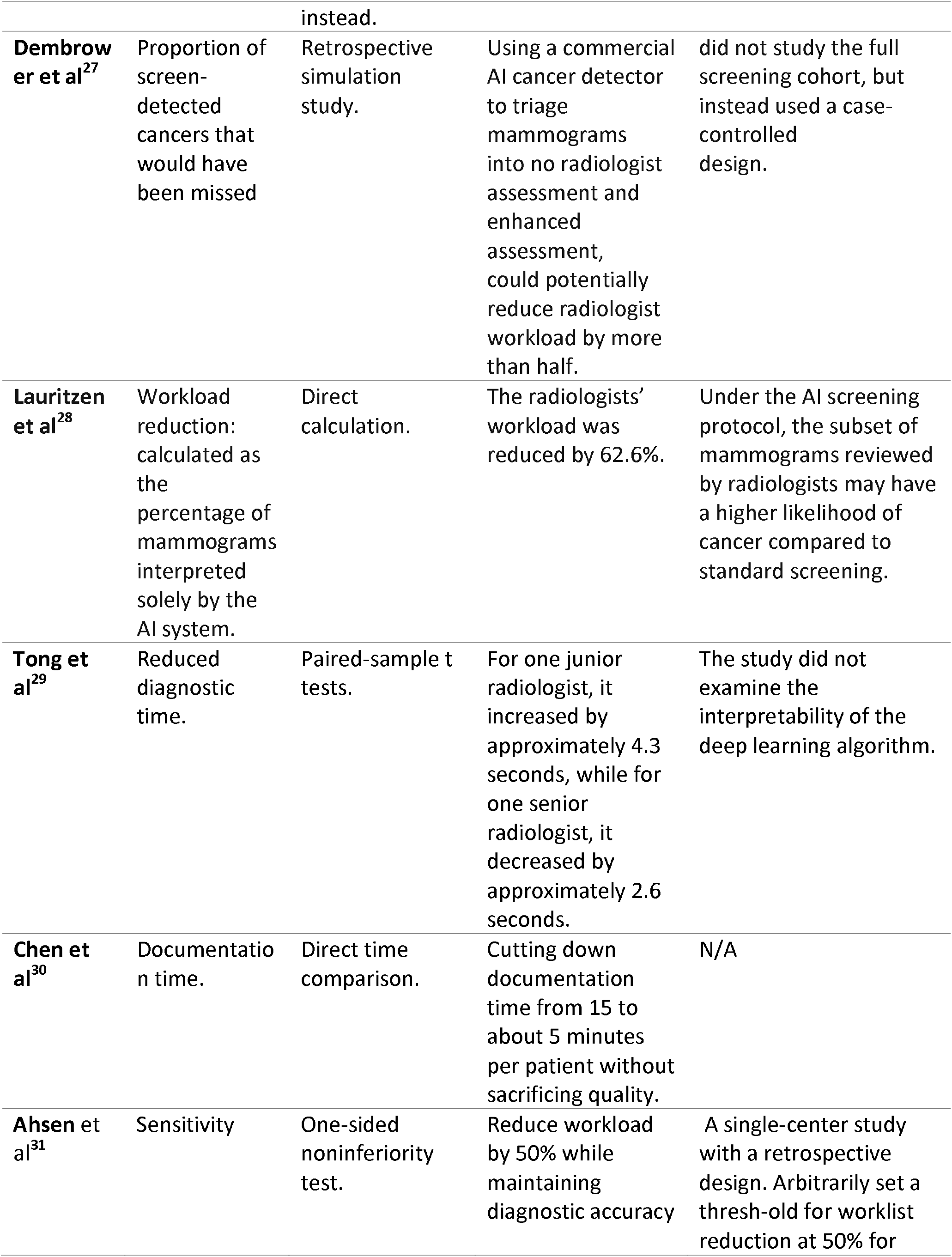
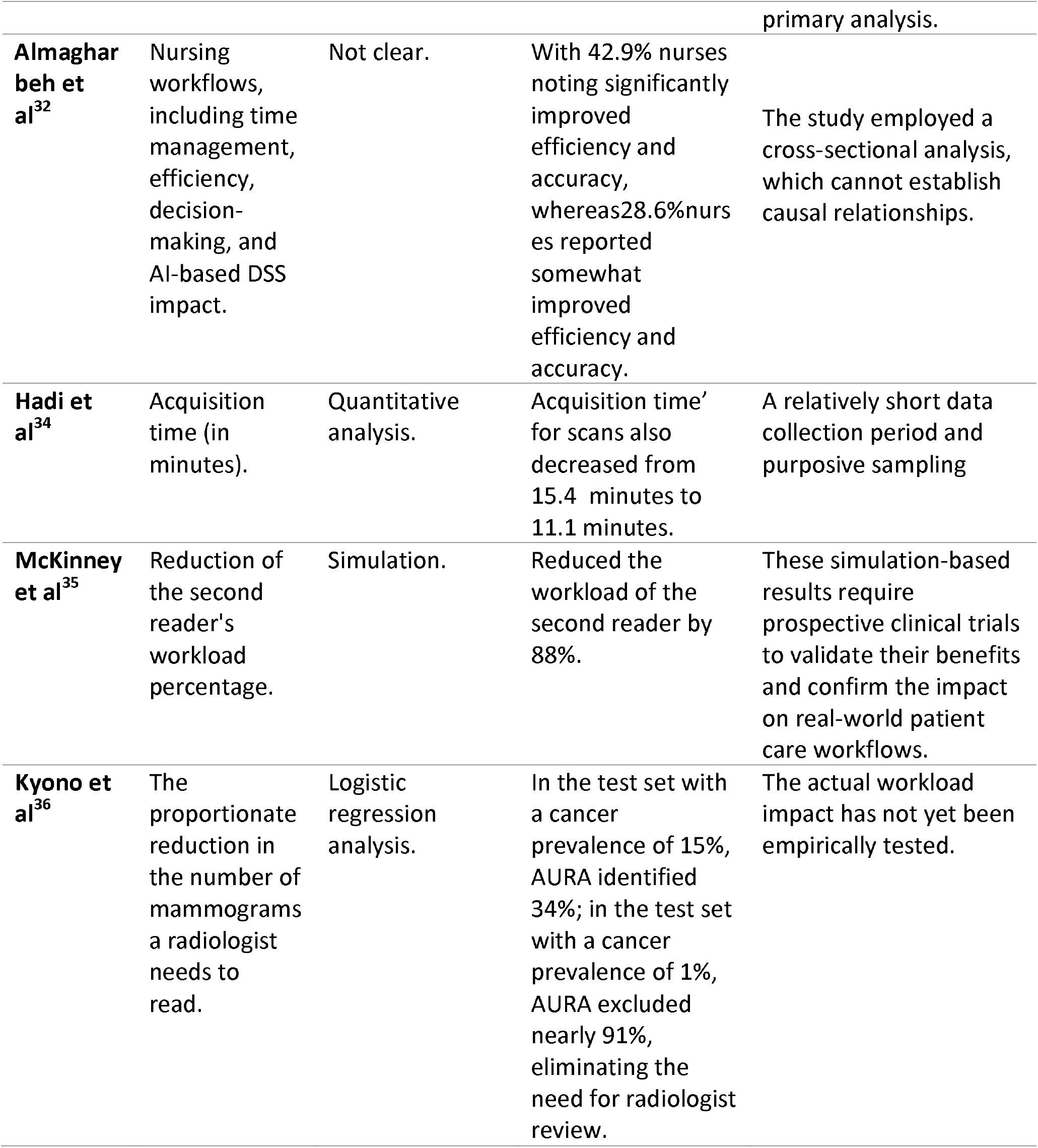
Methodological Approaches for Examining Workforce Productivity.

#### Analytical Approaches and Statistical Methods

The studies included in this review employed a diverse range of analytical approaches to estimate the impact of AI on healthcare costs, productivity, and efficiency. At the broader economic level, macroeconomic modeling and bottom-up use-case analysis were used to forecast potential system-level savings. For instance, Sahni et al.^19^ estimate that wider adoption of AI could lead to savings of 5% to 10% in U.S. healthcare spending—approximately $200 to $360 billion annually in 2019 dollars. These estimates are based on specific AI-enabled use cases that utilize current technologies and are attainable within the next five years without compromising quality or access. Their approach involved mapping specific healthcare tasks (e.g., administrative work, diagnostics, operations support) to AI capabilities and estimating time and cost reductions from AI adoption across sectors like payer services and hospital operations. This analysis offers a macroeconomic view of how AI could reshape healthcare spending patterns if deployed at scale.

In contrast, scenario-based cost modeling has also been applied to specific clinical workflows. Kacew et al.^14^ evaluated eight diagnostic strategies for determining mismatch repair/microsatellite instability (MMR/MSI) status in metastatic colorectal cancer. Their analysis found that a strategy combining high-sensitivity AI with confirmatory high-specificity PCR or IHC testing resulted in the greatest cost savings—approximately $400 million (12.9%)—compared to next-generation sequencing alone. Additionally, a high-specificity AI-only approach provided the shortest time to treatment initiation (<1 day) and maintained a 97% rate of patients receiving guideline-supported therapy.

Experimental and quasi-experimental designs were also employed to directly quantify changes in healthcare productivity resulting from AI use. A notable example is the cluster-randomized trial by Abramoff et al.^21^, which examined the use of an autonomous AI system for diabetic eye exams in primary care. The study used queueing theory to model clinic workflows and measured productivity by the number of completed patient encounters per hour. Clinics using the AI system achieved a 40% increase in throughput compared to controls, highlighting the operational gains achievable through autonomous decision-making tools.

## DISCUSSION

This review synthesizes current evidence on the macro-level effects of artificial intelligence (AI) across three critical domains of health system performance: the health economy, workforce productivity, and administrative efficiency. Across these domains, AI—particularly in the form of machine learning algorithms, generative models, and natural language processing systems— demonstrates substantial promise in generating cost savings, enhancing operational throughput, and improving workflow efficiency.

In the economic domain, analyses from sources such as McKinsey and the National Bureau of Economic Research estimate that scaled AI implementation could reduce national health expenditures by 5 to 10 percent. These projected savings are primarily attributed to automation of clinical and administrative processes and the deployment of predictive analytics. However, these forecasts are based on optimistic assumptions, and their applicability across healthcare systems with varying regulatory and infrastructural contexts remains uncertain.

AI has also shown promise in mitigating workforce inefficiencies. Applications such as ambient AI and large language models are increasingly used to automate clinical documentation and accelerate diagnostic workflows. Systematic reviews have reported significant reductions in diagnostic time—occasionally exceeding 90 percent. However, such gains are offset by ongoing concerns about the accuracy, reliability, and contextual coherence of AI-generated content, particularly in documentation where clinical nuance and narrative detail remain essential.

Administrative functions—including claims processing, prior authorization, and provider credentialing—emerge as particularly well-suited to AI-enabled automation. These high-cost, low-clinical-risk tasks present opportunities for efficiency, standardization, and transparency, with relatively lower ethical and safety concerns. As such, they may serve as practical entry points for broader AI integration at the system level.

### Limitations of Existing Studies

While the research on AI in healthcare is growing fast, there are still some major gaps we need to talk about. First, a lot of the studies are short-term, so we don’t really know how AI tools perform over the long run or how they change actual patient outcomes. Also, many of them rely on very specific patient groups or are based in just one or two hospitals, which means the findings might not apply to everyone. Another big issue is that most studies don’t break down their results by race, income level, or social background, so we’re missing a clear picture of whether AI is helping or hurting when it comes to health equity. On top of that, some studies struggle with confounding factors—like whether certain types of patients are more likely to get AI-assisted care—which can muddy the results. And finally, while we know that introducing AI can shake up how people work, very few papers look into the real-world barriers to getting these tools adopted, such as whether clinicians trust the tech or how it fits into their daily routines. So even though the potential is huge, there’s still a lot we don’t fully understand.

Despite the potential of artificial intelligence to enhance efficiency and reduce costs in healthcare, its unintended consequences warrant careful consideration. Overreliance on AI may erode clinicians’ diagnostic judgment, leading to skill degradation, increased risk of misdiagnosis, and the resulting costs of medical errors and retraining^39,40^. If training data used for AI models is biased, the technology may deliver inequitable outcomes, disproportionately impacting minority groups and further exacerbating health disparities, with increased long-term costs due to preventable disease progression^41,42^. While AI can improve diagnostic sensitivity, it may also result in incidental findings and overtreatment, contributing to the waste of limited healthcare resources. Furthermore, the vast amount of health data required by AI systems poses significant privacy and cybersecurity risks; data breaches may lead to legal liabilities and high infrastructure costs, especially burdensome for smaller institutions^43,44^. Although AI can alleviate workforce burdens, it may disrupt labor markets by displacing certain roles and creating reskilling needs. Highly specialized algorithms may also fragment care delivery, reducing coordination and integration. To mitigate these risks, the implementation of AI must be grounded in careful governance and equitable model design to ensure that it strengthens— rather than undermines—high-quality, patient-centered care^45,46^.

### Research and Policy Implications

Artificial intelligence is speeding image reading, documentation, and administrative workflows, but supporting evidence remains scattered across small pilots, heterogeneous designs, and simulations, hindering comparison and generalization^47 48^. Future studies should adopt shared efficiency metrics—time saved per case, full cost-utility ratios that include upkeep, and indicators such as user trust and alert overrides—while embedding prospective mixed-methods evaluations in routine care to link data quality, culture, and interface design with short-term performance and long-term sustainability^49^. Economic work must count real deployment costs—retraining, integration, workforce preparation—so decision-makers see net value, not idealized savings^47,50^.

It’s worth to note that efficiency gains alone will not necessarily ease shortages: while in nursing, AI may enhance nurses’ work-life balance^51^, some physicians are worried that they may have to see more patients in the case^52^. Large-scale deployments therefore need labor-market impact assessments that shape staffing benchmarks, reskilling programs, and wage structures, keeping progress humane^38^. Because data distributions shift—dramatically so during events like COVID-19—models need continual calibration and monitoring^53^; governance should mandate drift-detection dashboards, external validation, and agile updates^48^.

On top of that, early studies already flag automation bias. Prinster□let□lal^54^. showed feature-based explanations improve accuracy yet foster faster, less critical acceptance, while Patil□let□lal^55^. warn that unquestioned trust may heighten burnout and liability. Both were short term; longitudinal work must test whether routine exposure blunts clinicians ‘ability to spot hallucinations or edge-case errors.

Before nationwide deployment, small and representative pilots should precede national roll-outs, revealing how diverse users weigh AI advice and guiding interface refinements. Budgets must fund these cycles of testing, training, recalibration, and monitoring^50^, acknowledging hidden implementation costs and AI’s lifelong upkeep. To close evidence gaps, stepped-wedge cluster trials^56^, difference-in-differences analyses^57^, and sequential mixed-methods studies—supplemented by long-term drift-monitoring cohorts—may yield causal, scalable insights while capturing behavioral adaptations and safety signals. Paired with transparent cost accounting and workforce modules, such designs will turn scattered demonstrations into a robust evidence base.

Current evidence on economics of AI is constrained by methodological heterogeneity, limited use of real-world implementation data, and a predominant reliance on theoretical and algorithmic models. While projections suggest substantial savings and operational gains, these benefits remain contingent on how AI is integrated within complex clinical environments that allow widespread adoption of the technology. Policy strategies that align regulatory safeguards with practical implementation challenges—while addressing trust, data governance, and equity—will be central to realizing the sustainable value of AI in healthcare.

## METHODS

This study employed a systematic review methodology to synthesize existing literature on the macro-level impact of AI on healthcare systems, with a specific focus on health system costs and spending, workforce productivity, and administrative efficiency. Unlike systematic reviews, the systematic review approach allows for the integration of diverse types of evidence, making it particularly well-suited to emerging and multidisciplinary topics such as AI-driven health system reform.illustrates the conceptual framework guiding this review. The framework positions AI as a system-xlevel catalyst capable of generating gains across three interrelated domains: health economy, workforce productivity, and administrative efficiency. The overlapping nature of these gears reflects the interconnected effects of AI interventions, in which improvements in one area (e.g., administrative automation) may drive ripple effects in overall cost savings and labor optimization.

A comprehensive literature search was conducted using four major databases: PubMed, and Google Scholar. The search covered literature published between January 2020 and July 28 2025. Keywords and Medical Subject Headings (MeSH) were selected to reflect the core themes of the study, including terms related to artificial intelligence (such as “AI,” “machine learning,” “natural language processing,” “Neural Networks,” “Computer-Assisted Diagnosis,” “Decision Support Systems, Clinical,” “digital health,” “artificial intelligence-assisted,” “artificial intelligence-based,” “artificial intelligence-aided,” “deep learning,” “neural networks,” “AI intervention s,” “clinical decision support,” “computer aided,” and “predictive analytics”), health system economics (including “health expenditure,” “healthcare cost,” “Cost-Benefit Analysis,” “Cost Savings,” “cost analysis,” “economic impact,” and “health care cost reductions”), workforce and productivity (such as “workforce,” “workload,” “productivity,” “workload reduction,” “labor productivity,” “health systems,” “health systems,” interpretation times and ““clinician workload”), and administrative efficiency (including “Efficiency, Organizational,” “administrative efficiency,” “Workflow,” “documentation time,” “claims processing,” “reporting times,” “administrative burden,” and “workflow efficiency”). Boolean operators were used to structure the search and connect concept clusters, allowing for the inclusion of studies that explored intersections such as AI and health spending or AI and administrative efficiency.

Studies were eligible for inclusion if they examined the macro-level impact of AI on healthcare systems and reported outcomes related to total cost, national or regional health expenditure, workforce productivity, or the efficiency of administrative processes. Eligible studies employed empirical methods, simulation models, structured reviews, or policy analysis and were published in peer-reviewed journals or as policy reports from reputable health or economic institutions. Studies were excluded if they focused solely on patient- or clinic-level cost-effectiveness or cost-utility analyses; if they discussed only clinical outcomes without broader system-level implications; if they presented algorithm development without application to system performance; if they lacked clear data or methodological frameworks; or if they were non–peer-reviewed publications such as commentaries, editorials, or opinion pieces.

Given the systematic structure of this review, the synthesis of findings was conducted thematically, guided by the three pre-specified domains: health system costs and macro-level spending, workforce productivity and labor restructuring, and administrative efficiency and system process optimization. The analysis also aimed to identify recurring themes, gaps in the evidence, methodological challenges, and insights specific to regional or health system contexts. For each included study, relevant information was extracted, including the country or region of study, the domain of AI application, study design, outcome measures, and key findings related to cost, productivity, or administrative efficiency. A flow diagram of records found, screened, selected, and excluded with corresponding exclusion criteria is shown in Figure 2. The remaining 24 studies were read full text.

**Figure 2.**
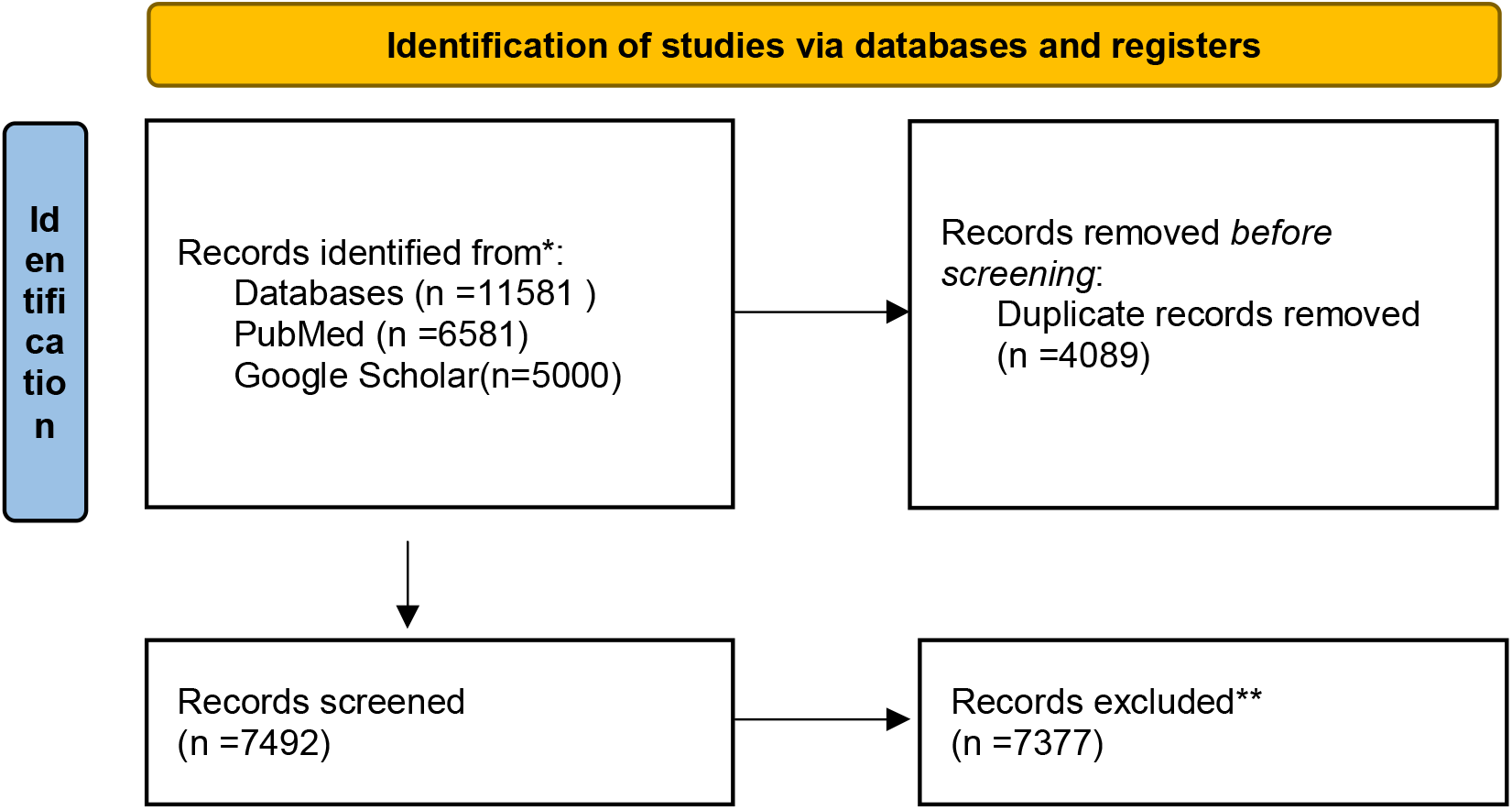
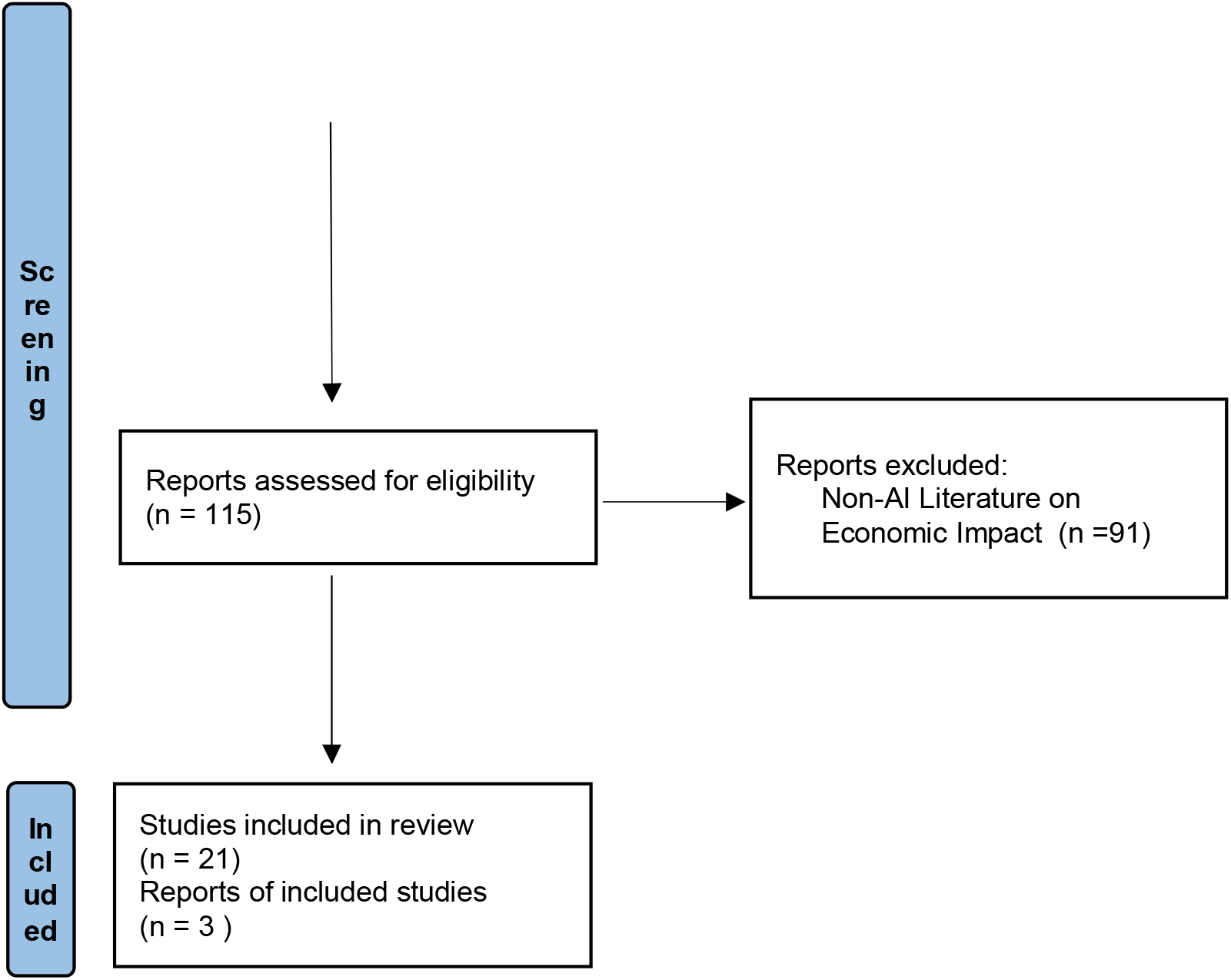
PRISMA flowchart describing study selection and reasons for exclusion during full-text screening.

## Data Availability

No data is used or generated in this systematic review

## Acknowledgements

We thank the Yushan Fellow Program by the Ministry of Education (MOE), Taiwan for the financial support. (MOE-112-YSFMN-0003-002-P1)

## Author contributions

J.T.L. conceived the study. J.T.L, V.T.L and wrote the first draft of the manuscript. V.T.N.L., V.C.S.L., C.W.H. and H.H.C. reviewed the article. All authors contributed to writing the second draft and have read and approved the final manuscript.

## Notes

### Competing Interest Statement

The authors have declared no competing interest.

### Funding Statement

This study is funded by Yushan Fellow Program from the Ministry of Education (MOE), Taiwan (MOE-112-YSFMN-0003-002-P1)

## REFERENCE

1. Trends in future health financing and coverage: future health spending and universal health coverage in 188 countries, 2016-40. Lancet. May 5 2018;391(10132):1783–1798. doi:10.1016/s0140-6736(18)30697-4

2. Fiore JA, Madison AJ, Poisal JA, et al. National Health Expenditure Projections, 2023-32: Payer Trends Diverge As Pandemic-Related Policies Fade. Health Aff (Millwood). Jul 2024;43(7):910–921. doi:10.1377/hlthaff.2024.00469

3. Past, present, and future of global health financing: a review of development assistance, government, out-of-pocket, and other private spending on health for 195 countries, 1995-2050. Lancet. Jun 1 2019;393(10187):2233–2260. doi:10.1016/s0140-6736(19)30841-4

4. Gainsbury S. How much more money does the NHS need? https://www.nuffieldtrust.org.uk/news-item/how-much-more-money-does-the-nhs-need

5. Elizabeth Williams AB, and Robin Rudowitz. Putting $880 Billion in Potential Federal Medicaid Cuts in Context of State Budgets and Coverage. KFF. https://www.kff.org/medicaid/issue-brief/putting-880-billion-in-potential-federal-medicaid-cuts-in-context-of-state-budgets-and-coverage/

6. Lee JT, Crettenden I, Tran M, et al. Methods for health workforce projection model: systematic review and recommended good practice reporting guideline. Hum Resour Health. Apr 17 2024;22(1):25. doi:10.1186/s12960-024-00895-z

7. Sinsky C, Colligan L, Li L, et al. Allocation of Physician Time in Ambulatory Practice: A Time and Motion Study in 4 Specialties. Ann Intern Med. Dec 6 2016;165(11):753–760. doi:10.7326/m16-0961

8. Toscano F, O’Donnell E, Broderick JE, et al. How Physicians Spend Their Work Time: an Ecological Momentary Assessment. J Gen Intern Med. Nov 2020;35(11):3166–3172. doi:10.1007/s11606-020-06087-4

9. Topol EJ. As artificial intelligence goes multimodal, medical applications multiply. Science. Sep 15 2023;381(6663):adk6139. doi:10.1126/science.adk6139

10. Moura L, Jones DT, Sheikh IS, et al. Implications of Large Language Models for Quality and Efficiency of Neurologic Care: Emerging Issues in Neurology. Neurology. Jun 11 2024;102(11):e209497. doi:10.1212/wnl.0000000000209497

11. Vo V, Chen G, Aquino YSJ, Carter SM, Do QN, Woode ME. Multi-stakeholder preferences for the use of artificial intelligence in healthcare: A systematic review and thematic analysis. Soc Sci Med. Dec 2023;338:116357. doi:10.1016/j.socscimed.2023.116357

12. Natesan D, Eisenstein EL, Thomas SM, et al. Health Care Cost Reductions with Machine Learning-Directed Evaluations during Radiation Therapy - An Economic Analysis of a Randomized Controlled Study. Nejm ai. Apr 2024;1(4)doi:10.1056/aioa2300118

13. Lavoie-Gagne O, Woo JJ, Williams RJ, 3rd, Nwachukwu BU, Kunze KN, Ramkumar PN. Artificial Intelligence as a Tool to Mitigate Administrative Burden, Optimize Billing, Reduce Insurance- and Credentialing-Related Expenses, and Improve Quality Assurance Within Health Care Systems. Arthroscopy. Mar 20 2025;doi:10.1016/j.arthro.2025.02.038

14. Kacew AJ, Strohbehn GW, Saulsberry L, et al. Artificial Intelligence Can Cut Costs While Maintaining Accuracy in Colorectal Cancer Genotyping. Front Oncol. 2021;11:630953. doi:10.3389/fonc.2021.630953

15. Yacoub B, Varga-Szemes A, Schoepf UJ, et al. Impact of Artificial Intelligence Assistance on Chest CT Interpretation Times: A Prospective Randomized Study. AJR Am J Roentgenol. Nov 2022;219(5):743–751. doi:10.2214/ajr.22.27598

16. Hunter JG, Bera K, Shah N, et al. Real-World Performance of Pneumothorax-Detecting Artificial Intelligence Algorithm and its Impact on Radiologist Reporting Times. Acad Radiol. Mar 2025;32(3):1165–1174. doi:10.1016/j.acra.2024.10.012

17. Chui M, Hazan E, Roberts R, et al. The economic potential of generative AI. 2023. McKinsey & Company. June.

18. Company M. Transforming healthcare with AI:The impact on the workforce and organisations. 2020. McKinsey & Company. March.

19. Sahni N, Stein G, Zemmel R, Cutler DM. THE POTENTIAL IMPACT OF ARTIFICIAL INTELLIGENCE ON HEALTHCARE SPENDING. 2023. http://www.nber.org/papers/w30857

20. Mori Y, Kudo SE, East JE, et al. Cost savings in colonoscopy with artificial intelligence-aided polyp diagnosis: an add-on analysis of a clinical trial (with video). Gastrointest Endosc. Oct 2020;92(4):905–911 e1. doi:10.1016/j.gie.2020.03.3759

21. Abramoff MD, Whitestone N, Patnaik JL, et al. Autonomous artificial intelligence increases real-world specialist clinic productivity in a cluster-randomized trial. NPJ Digit Med. Oct 4 2023;6(1):184. doi:10.1038/s41746-023-00931-7

22. Worrall C, Shirley D, Bullard J, Dao A, Morrisette T. Impact of a clinical pharmacist-led, artificial intelligence-supported medication adherence program on medication adherence performance, chronic disease control measures, and cost savings. J Am Pharm Assoc (2003). Jan-Feb 2025;65(1):102271. doi:10.1016/j.japh.2024.102271

23. Chen M, Wang Y, Wang Q, et al. Impact of human and artificial intelligence collaboration on workload reduction in medical image interpretation. NPJ Digit Med. Nov 30 2024;7(1):349. doi:10.1038/s41746-024-01328-w

24. Willmen T, Völkel L, Ronicke S, et al. Health economic benefits through the use of diagnostic support systems and expert knowledge. BMC Health Serv Res. Sep 9 2021;21(1):947. doi:10.1186/s12913-021-06926-y

25. Brix MAK, Järvinen J, Bode MK, et al. Financial impact of incorporating deep learning reconstruction into magnetic resonance imaging routine. Eur J Radiol. Jun 2024;175:111434. doi:10.1016/j.ejrad.2024.111434

26. Rozario N, Rozario D. Can machine learning optimize the efficiency of the operating room in the era of COVID-19? Can J Surg. Nov-Dec 2020;63(6):E527–e529. doi:10.1503/cjs.016520

27. Dembrower K, Wåhlin E, Liu Y, et al. Effect of artificial intelligence-based triaging of breast cancer screening mammograms on cancer detection and radiologist workload: a retrospective simulation study. Lancet Digit Health. Sep 2020;2(9):e468–e474. doi:10.1016/s2589-7500(20)30185-0

28. Lauritzen AD, Rodríguez-Ruiz A, von Euler-Chelpin MC, et al. An Artificial Intelligence-based Mammography Screening Protocol for Breast Cancer: Outcome and Radiologist Workload. Radiology. Jul 2022;304(1):41–49. doi:10.1148/radiol.210948

29. Tong WJ, Wu SH, Cheng MQ, et al. Integration of Artificial Intelligence Decision Aids to Reduce Workload and Enhance Efficiency in Thyroid Nodule Management. JAMA Netw Open. May 1 2023;6(5):e2313674. doi:10.1001/jamanetworkopen.2023.13674

30. Chen CJ, Liao CT, Tung YC, Liu CF. Enhancing Healthcare Efficiency: Integrating ChatGPT in Nursing Documentation. Stud Health Technol Inform. Aug 22 2024;316:851–852. doi:10.3233/shti240545

31. Ahsen ME, Ayvaci MUS, Mookerjee R, Stolovitzky G. Economics of AI and human task sharing for decision making in screening mammography. Nat Commun. Mar 7 2025;16(1):2289. doi:10.1038/s41467-025-57409-1

32. Almagharbeh WT. The impact of AI-based decision support systems on nursing workflows in critical care units. Int Nurs Rev. Jun 2025;72(2):e13011. doi:10.1111/inr.13011

33. Evans K, Papinniemi A, Ploderer B, et al. Impact of using an AI scribe on clinical documentation and clinician-patient interactions in allied health private practice: perspectives of clinicians and patients. Musculoskelet Sci Pract. Aug 2025;78:103333. doi:10.1016/j.msksp.2025.103333

34. Hadi YH, Altalhi FK, Ali HM, Shabli MA, Aqil AIA, England A. Enhancing CT examination efficiency with ChatGPT-4o for multilingual Hajj pilgrims: A short communication. J Med Imaging Radiat Sci. Jan 2025;56(1):101781. doi:10.1016/j.jmir.2024.101781

35. McKinney SM, Sieniek M, Godbole V, et al. International evaluation of an AI system for breast cancer screening. Nature. Jan 2020;577(7788):89–94. doi:10.1038/s41586-019-1799-6

36. Kyono T, Gilbert FJ, van der Schaar M. Improving Workflow Efficiency for Mammography Using Machine Learning. J Am Coll Radiol. Jan 2020;17(1 Pt A):56–63. doi:10.1016/j.jacr.2019.05.012

37. Dvijotham KD, Winkens J, Barsbey M, et al. Enhancing the reliability and accuracy of AI-enabled diagnosis via complementarity-driven deferral to clinicians. Nat Med. Jul 2023;29(7):1814–1820. doi:10.1038/s41591-023-02437-x

38. Professor Sandeep Reddy C, Centre for Advancement of Translational AI in Medicine & Member, Professional Development Committee, HIMSS. The Impact of AI on the Healthcare Workforce: Balancing Opportunities and Challenges. https://legacy.himss.org/resources/impact-ai-healthcare-workforce-balancing-opportunities-and-challenges?utm_source=chatgpt.com

39. Ferryman K, Mackintosh M, Ghassemi M. Considering Biased Data as Informative Artifacts in AI-Assisted Health Care. N Engl J Med. Aug 31 2023;389(9):833–838. doi:10.1056/NEJMra2214964

40. Seyyed-Kalantari L, Zhang H, McDermott MBA, Chen IY, Ghassemi M. Underdiagnosis bias of artificial intelligence algorithms applied to chest radiographs in under-served patient populations. Nat Med. Dec 2021;27(12):2176–2182. doi:10.1038/s41591-021-01595-0

41. Siddique SM, Tipton K, Leas B, et al. The Impact of Health Care Algorithms on Racial and Ethnic Disparities : A Systematic Review. Ann Intern Med. Apr 2024;177(4):484–496. doi:10.7326/m23-2960

42. Armoundas AA, Narayan SM, Arnett DK, et al. Use of Artificial Intelligence in Improving Outcomes in Heart Disease: A Scientific Statement From the American Heart Association. Circulation. Apr 2 2024;149(14):e1028–e1050. doi:10.1161/cir.0000000000001201

43. Haug CJ, Drazen JM. Artificial Intelligence and Machine Learning in Clinical Medicine, 2023. N Engl J Med. Mar 30 2023;388(13):1201–1208. doi:10.1056/NEJMra2302038

44. Senkaiahliyan S, Detsky AS, Ma J, Lawler PR, Paramasamy J, Visser JJ. The Economic, Political, and Societal Consequences of Disrupting Health Care Delivery with Artificial Intelligence. J Gen Intern Med. May 8 2025;doi:10.1007/s11606-025-09590-8

45. Occhipinti JA, Prodan A, Hynes W, et al. Artificial intelligence, recessionary pressures and population health. Bull World Health Organ. Feb 1 2025;103(2):155–163. doi:10.2471/blt.24.291950

46. Esmaeilzadeh P. Challenges and strategies for wide-scale artificial intelligence (AI) deployment in healthcare practices: A perspective for healthcare organizations. Artif Intell Med. May 2024;151:102861. doi:10.1016/j.artmed.2024.102861

47. Khanna NN, Maindarkar MA, Viswanathan V, et al. Economics of Artificial Intelligence in Healthcare: Diagnosis vs. Treatment. Healthcare (Basel). Dec 9 2022;10(12)doi:10.3390/healthcare10122493

48. Chan HP, Samala RK, Hadjiiski LM, Zhou C. Deep Learning in Medical Image Analysis. Adv Exp Med Biol. 2020;1213:3–21. doi:10.1007/978-3-030-33128-3_1

49. Andersen ES, Birk-Korch JB, Röttger R, Brasen CL, Brandslund I, Madsen JS. Monitoring performance of clinical artificial intelligence: a scoping review protocol. JBI Evid Synth. Mar 1 2024;22(3):453–460. doi:10.11124/jbies-23-00390

50. Bajwa J, Munir U, Nori A, Williams B. Artificial intelligence in healthcare: transforming the practice of medicine. Future Healthc J. Jul 2021;8(2):e188–e194. doi:10.7861/fhj.2021-009551.

51. Rony MKK, Alrazeeni DM, Akter F, et al. The role of artificial intelligence in enhancing nurses’ work-life balance. Journal of Medicine, Surgery, and Public Health. 2024;3doi:10.1016/j.glmedi.2024.100135

52. Bundy H, Gerhart J, Baek S, et al. Can the Administrative Loads of Physicians be Alleviated by AI-Facilitated Clinical Documentation? J Gen Intern Med. Nov 2024;39(15):2995–3000. doi:10.1007/s11606-024-08870-z

53. Sahiner B, Chen W, Samala RK, Petrick N. Data drift in medical machine learning: implications and potential remedies. Br J Radiol. Oct 2023;96(1150):20220878. doi:10.1259/bjr.20220878

54. Prinster D, Mahmood A, Saria S, et al. Care to Explain? AI Explanation Types Differentially Impact Chest Radiograph Diagnostic Performance and Physician Trust in AI. Radiology. Nov 2024;313(2):e233261. doi:10.1148/radiol.233261

55. Patil SV, Myers CG, Lu-Myers Y. Calibrating AI Reliance-A Physician’s Superhuman Dilemma. JAMA Health Forum. Mar 7 2025;6(3):e250106. doi:10.1001/jamahealthforum.2025.0106

56. Heagerty PJMD. Experimental Designs and Randomization Schemes: Stepped-Wedge Designs. https://rethinkingclinicaltrials.org/chapters/design/experimental-designs-and-randomization-schemes/stepped-wedge-designs/

57. Fredriksson Aao, Gustavo. Impact evaluation using Difference-in-Differences. RAUSP management journa. 2019;doi:10.1108/RAUSP-05-2019-0112

